# Diagnostic performance of the Pluslife MiniDock MTB and Molbio MTB Ultima assays to detect tuberculosis from tongue and sputum swabs among outpatients and in active case finding in Cameroon

**DOI:** 10.1101/2025.09.11.25335435

**Authors:** Cyrille Mbuli, Taya Fokou Jean Bosco, Rita Nsamenang, Biatu Nestor, Gildas Nguimfack, Ngha Ndze Mbuh, Joceline Konso, Zourriyah Adamou Mana, Nankouo Arthur, Tiamuh Nzebele Magdalene, Nsame Denis, Norah Nyah Ndi, Irene Adeline Goupeyou Wandji, Mercy Fundoh, Maurice Ganava Toussaint, Ousmanou Bello, Meoto Paul, Fitame Adeline, Vuchas Comfort, Pride Teyim, Valerie Flore Donkeng Donfack, Tushar Garg, Jacob Creswell, Melissa Sander, the RAPID TB Consortium

**Affiliations:** Center for Health Promotion and Research, Bamenda, Cameroon; Bamenda Regional Hospital, Bamenda, Cameroon; Cameroon Baptist Convention Health Services, Bamenda, Cameroon; National TB Program – Littoral Region, Douala, Cameroon; National TB Program – Northwest Region, Bamenda, Cameroon; National TB Program – Far North Region, Maroua, Cameroon; National TB Program – North Region, Garoua, Cameroon; National TB Program – Southwest Region, Buea, Cameroon; National TB Program – West Region, Bafoussam, Cameroon; Tuberculosis Reference Laboratory Douala, Douala, Cameroon; Centre Pasteur du Cameroun, Yaoundé, Center, Cameroun; Stop TB Partnership, Geneva, Switzerland

**Keywords:** tuberculosis, diagnostic

## Abstract

**Background:** Cost and infrastructure requirements limit access to current rapid molecular diagnostic testing for tuberculosis (TB). Recent developments in novel, swab-based assays that can be used closer to the point of care offer the potential to change the TB diagnostic landscape.

**Methods:** We assessed the diagnostic accuracy of two tests, Molbio Truenat MTB Ultima (MTB Ultima) and Pluslife MiniDock MTB Test (MiniDock MTB), among consecutively enrolled individuals aged 15 and above in health facilities and community-based screening events in Cameroon, compared to the reference standard of TB culture. Two tongue swabs and sputum specimens were requested from each participant. Comparator tests were smear microscopy and Xpert MTB/RIF Ultra.

**Results:** From February to June 2025, 1,097 participants were enrolled in communities (382) and at health facilities (715). Sensitivities of sputum and tongue swabs on MiniDock MTB among 132 people with culture-positive TB were 86% (95% CI, 79-91%) and 76% (95% CI, 68-82%), respectively, and 67% (95%CI, 51-79%) and 44% (95%CI, 29-59%) among those with smear-negative TB. Sensitivities of sputum and tongue swabs on MTB Ultima were 84% (97/116, 95%CI, 76-89%) and 74% (67/91, 95% CI, 64-82%), respectively, and 58% (95% CI, 41-74%) and 33% (95% CI, 18-53%) among those with smear-negative TB. Specificities of both tests were high (>97%).

**Conclusions:** In this population, the performance of both MiniDock MTB and MTB Ultima on tongue and sputum swabs was similar to target product profile thresholds for near point of care TB tests. Further studies to evaluate performance in diverse populations and settings are needed.

**Summary:** Our diagnostic accuracy study of new swab-based MiniDock MTB and MTB Ultima assays in Cameroon demonstrated accuracies similar to the tuberculosis detection target product profiles overall for near point of care tongue and sputum swabs.

## Introduction

Tuberculosis (TB) kills more people globally (1.25 million in 2023) than any other infectious disease, more than HIV and malaria combined[1]. This is largely due to the roughly 3 million people with incident TB missed by health systems each year alongside millions more missed with prevalent disease[1].

Sputum microscopy had been the cornerstone of TB diagnosis for decades, but has limited sensitivity, especially among people living with HIV and children, and is highly dependent on human performance[2]. In 2010, the introduction of Cepheid’s (Sunnyvale, USA) GeneXpert system and Xpert MTB/RIF (Xpert) cartridge-based automated nucleic acid amplification test (NAAT) promised to revolutionize TB diagnostics[3]. While improvements in TB notifications were not as significant as hoped due to associated drops in clinical diagnosis, bacteriological confirmation improved[4]. In 2016, a manual NAAT platform, the Loopamp MTBC assay (TB LAMP, Eiken, Japan), was introduced as a lower-cost alternative for the detection of TB[5]. The next-generation Xpert MTB/RIF Ultra (Ultra) further improved sensitivity compared to the Xpert[6], and together with Molbio Diagnostics (Goa, India) Truenat platform[7,8], these NAAT have become the global standard for the detection of TB and rifampin resistance [9].

Unfortunately, despite two price reductions, molecular testing coverage remains unacceptably low. In 2023, less than half of people with TB received a rapid molecular assay as their initial diagnostic test[1,10]. This shortcoming can largely be attributed to cost and complexity. Even at current prices of just under USD 8, the NAATs have expensive modular components, maintenance and service add to real per test costs[11,12], and limited national budgets have been stretched even thinner by the US government’s recent cuts[13,14]. Additionally, the infrastructure needed to use the platforms has generally relegated current low-complexity NAAT devices to district level care, while many patient pathway analyses have shown that people with TB first seek care at lower levels in the healthcare system[15,16].

Building off investments made during the COVID-19 pandemic, several companies have developed simplified diagnostic platforms that can test oral swabs as well as swabbed sputum, and have the potential to be placed at lower levels of care to increase access to testing [17]. One such assay is Molbio MTB Ultima (Goa, India) with the new Truelyse device that replaces the Trueprep extraction device with a sonification lysis step, followed by real-time PCR on the Truelab instrument. The other is MiniDock MTB (Guangzhou Pluslife Biotech, China) with an initial mechanical lysis step (Thermomixer) and then the MiniDock instrument for amplification and detection[18]. Steadman et al. reported on the performance of these two platforms for tongue swabs and swabbed sputum in outpatients in India, Uganda, and Vietnam[19]. In the population tested, the sensitivity of both platforms was similar to Ultra on swabbed sputum and significantly better than microscopy on tongue swabs, with high specificities.

Additional evaluations of the performance of these new assays to detect TB are needed, in different populations and settings, including in the community. We report on a diagnostic accuracy study of both platforms among individuals enrolled in facility and community settings in Cameroon.

## Methods

### Study design

This was a prospective, multisite, diagnostic accuracy study of the performance of the MiniDock MTB and MTB Ultima assays conducted in Cameroon, at five health facilities and as part of active case finding in community health camps (Supplementary Table 1).

**Table 1.** Participant demographic and clinical characteristics, overall and by setting.

| Characteristic | Overall | Active case finding | Health facilities |
| --- | --- | --- | --- |
| Total | 1,097 | 382 | 715 |
| Age, years | 49 (36-63) | 52 (40-67) | 47 (34-60) |
| Female | 487 (44) | 157 (41) | 330 (46) |
| People living with HIV<sup>a</sup> | 205 (19) | 7 (2) | 198 (28) |
| People with diabetes | 88 (8) | 21 (5) | 67 (9) |
| Chest X-ray, AI score |  | 0.86 (0.62-0.96) |  |
| Cough > 2 weeks | 494 (45) | 143 (37) | 351 (49) |
| TB symptoms, any of W4SS | 920 (84) | 327 (86) | 593 (83) |
| Tongue swab collection method |  |  |  |
| Supervised self-collection | 915 (83) | 305 (80) | 610 (85) |
| HCW collection | 181 (16) | 77 (20) | 104 (15) |
| Sputum Ultra positive | 163 (15) | 57 (15) | 106 (15) |
| High | 61 (37) | 6 (11) | 55 (52) |
| Medium | 23 (14) | 6 (11) | 17 (16) |
| Low | 34 (21) | 18 (32) | 16 (15) |
| Very low | 19 (12) | 15 (26) | 4 (4) |
| Trace | 26 (16) | 12 (21) | 14 (13) |
| Sputum TB culture positive | 132 (12) | 32 (8) | 100 (14) |
| Sputum smear-positive<sup>b</sup> | 90 (8) | 19 (5) | 71 (10) |

n(%) or median(IQR); W4SS: WHO- recommended four symptom screen
<sup>a</sup>101 with HIV status unknown (all in ACF); <sup>b</sup>3 with smear result missing

At health facilities, consecutive outpatients aged 15 years and above with symptoms and/or clinical risk factors (diabetes, current smoker) for TB were invited to participate. At community-based active case finding (ACF) events, people aged 15 years and above were screened using chest X-ray with AI (qXR V4, Qure.ai, Mumbai, India), and those with an AI score of ≥0.30 were invited to participate. Individuals who were on TB treatment within the previous six months were excluded.

### Procedures

After providing written informed consent, participant demographic information and medical history were recorded. Two tongue swabs and two sputum specimens were requested from each participant (Figure 1).

**Figure 1.**
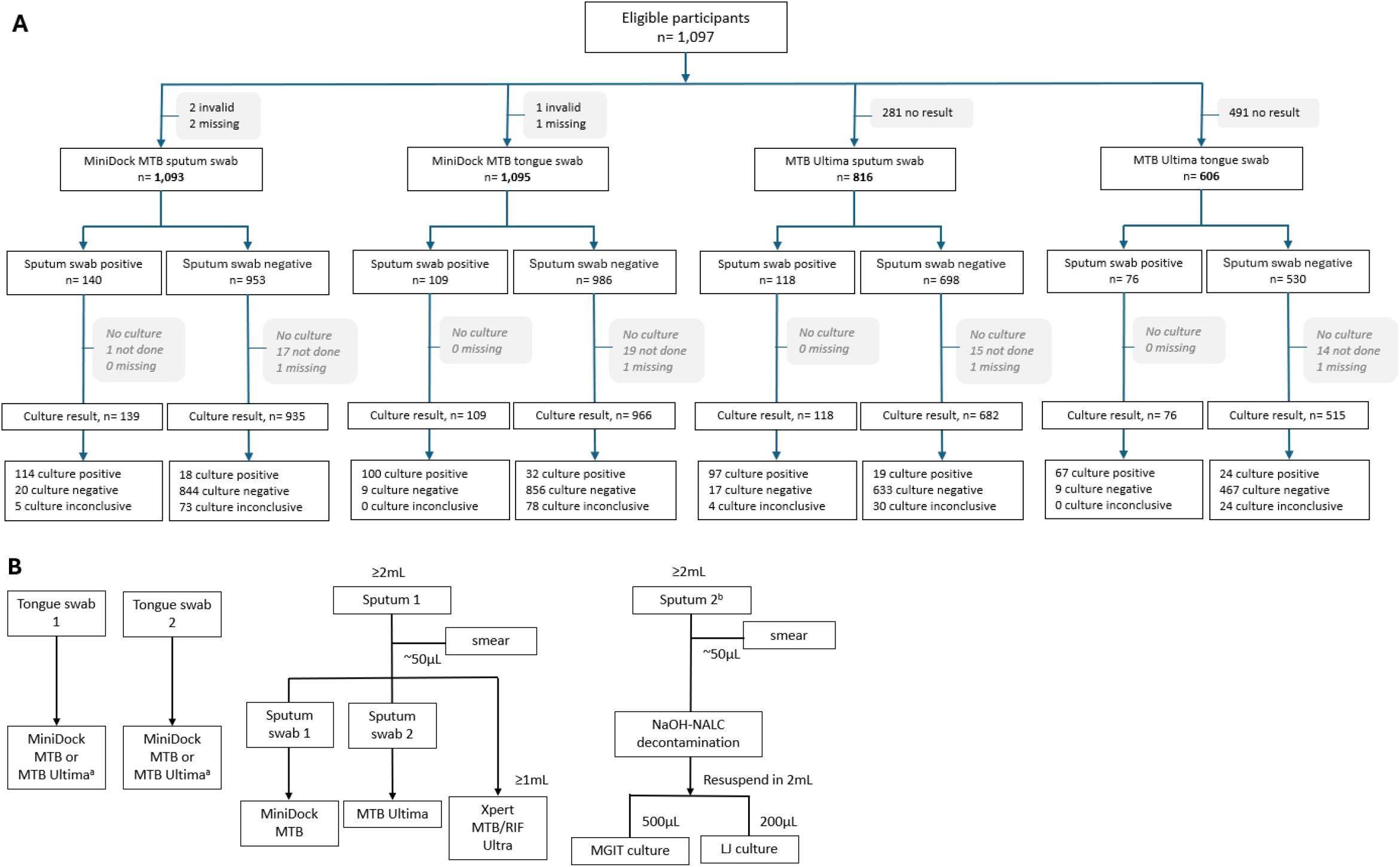
A, Participant flow. B, Specimen and testing flow. **a.** Tongue swabs were randomized after collection to either #1-MiniDock / #2-Ultima *or* #1-Ultima / #2-MiniDock; **b.** 2mL of sputum was processed for culture; specimens with >2mL were homogenized and excess was aliquoted and stored; at health facilities, two additional tongue swabs and a 3rd sputum specimen were collected on Day 2 when possible; stored sputum and pellets were used for additional testing in case of contaminated or discordant results.

### Specimen collection

Participants were shown a short video on how to self-collect tongue swabs, including swabbing the dorsum while rotating the swab for at least 30 seconds[20]. Then the participant was asked if they would self-collect a specimen under supervision; for participants who elected not to self-collect, a healthcare worker collected the tongue swabs. Swabs were collected, after ensuring that the participant had consumed nothing by mouth within 30 minutes, with nylon, flocked swabs (Copan FLOQSwabs 502CS01 or 520CS01) and stored dry in sterile tubes. Two swabs were collected successively; the testing sequence was randomized (Supplemental methods)[21].

Following tongue swab collection, participants were instructed on how to produce two sputum specimens with a volume of ≥2 mL each, collected at least 30 minutes apart. If the participant had difficulty to produce sputum, they were shown a video on sputum production and/or offered the Lung Flute ECO (Acoustic Innovations, Japan) with video coaching on how to use this device to assist sputum collection[22].

From each sputum specimen, a smear was prepared for microscopy, and two swabs were prepared by swirling the swab in sputum ten times over 10-15 seconds, then stored in sterile dry tubes until testing.

Swabs for MiniDock MTB were stored at room temperature and tested within 4 hours or stored at 2-8°C and tested within 24 hours; swabs for MTB Ultima were transferred to the reference lab and stored at -20°C for up to 5 months before testing. The second sputum specimen was stored at 2-8°C and transported to the reference laboratory for culture.

### Index tests

MiniDock MTB tests were performed onsite during 46 ACF events and in one of two laboratories (Supplemental Table 1). Tests were performed following manufacturer’s instructions. The MiniDock MTB provides results of positive, negative or invalid.

Ultima MTB tests were performed at the reference lab on stored specimens, which were first thawed to room temperature and then tested following the manufacturer’s instructions, with results from the TrueLab as MTB detected (high, medium, low, or very low), MTB not detected or invalid.

Invalid tests were repeated once, with the repeat result reported. Index tests were performed by technicians without knowledge of any other TB test results or clinical information.

### Comparator and reference tests

Smear microscopy was performed by Ziehl-Neelsen or fluorescence microscopy. The remainder of the first sputum was used for Ultra testing, following manufacturer’s instructions. The Xpert MTB/RIF Ultra assay produces results of invalid, error, MTB not detected, or MTB detected (high, medium, low, very low, or trace).

At the reference laboratory, sputum was processed for culture using the NaOH-NALC method and inoculated onto liquid and solid media (Supplemental methods) [23]. Technicians performing smear, Ultra and culture testing were blinded to index test results.

### Data analysis

The sample size was calculated as a minimum of 112 people with culture-positive TB, using a sensitivity target of 75% (based on the target product profile (TPP) for near-point-of-care, non-sputum specimens)[24] and a 95% confidence interval of +/-8%

For the diagnostic accuracy analysis, a microbiological reference standard based on culture from one sputum specimen was used, including one automated liquid culture and one solid culture (Figure 1). Participants for whom at least one culture was positive for TB were defined as having culture-positive TB; among these, anyone with at least one smear positive for TB was defined as smear positive.

Participants for whom MGIT or both cultures were negative for TB were defined as culture negative for TB. For the comparator, any Ultra result with MTB detected was considered positive. Participants were characterized using simple descriptive statistics. To evaluate the accuracy and agreement of the MiniDock MTB and MTB Ultima assays, we calculated the sensitivity and specificity point estimates with 95% confidence intervals (CIs) using the Wilson score method. We referred to guidance for conducting studies to evaluate the accuracy of sputum-based tests to detect TB for study design and reporting of results.[25]. The Standards for Reporting of Diagnostic Accuracy (STARD) recommendations were followed for reporting these data.[26] Analyses were performed in R version 4.5.1.

This study was approved by the National Ethics Committee for Research on Human Health and by the Institutional Review Board of the Cameroon Baptist Convention Health Board. This study was registered at PACTR (202502473140037). Study data were collected and managed using (Research Electronic Data Capture) REDCap tools.

## Results

### Participants

From February to June 2025, 1,097 participants were enrolled, with index and microbiological reference standard results as shown in Figure 1. Of these, 44% were female, the median age was 49 years, 19% were people known to be living with HIV, and 132 (12%) had culture-positive TB (Table 1). Ultra grade distribution differed by setting, with 47% of those in ACF having grades of very low or trace, as compared to 17% of outpatients. Overall, 1,095 (99.8%) provided a first sputum specimen, 1,077 provided two sputum specimens, and 1,096 (99.9%) provided two tongue swab specimens.

### MiniDock MTB diagnostic accuracy and agreement

For the MiniDock MTB, overall sensitivity of sputum and tongue swabs were 86% (95% CI, 79-91%) and 76% (95% CI, 68-82%), respectively, as compared to sputum Ultra of 95% (95% CI, 90-98%) (Table 2). Among smear-negative specimens, the sensitivity of sputum swabs was 67% (95% CI, 51-79%), of tongue swabs 44% (95% CI, 29-59%), as compared to sputum Ultra of 85% (95% CI, 70-93%).

**Table 2.** Diagnostic accuracy of tongue swabs and sputum swabs on MiniDock MTB and sputum on Xpert MTB/RIF Ultra for detection of TB, as compared with the reference standard of TB culture.

|  | N | Sensitivity: all culture positive, %<br>(95% CI, n/N) | Sensitivity: smear positive*, %<br>(95% CI, n/N) | Sensitivity: smear negative, %<br>(95% CI, n/N) | Specificity** %<br>(95% CI, n/N) |
| --- | --- | --- | --- | --- | --- |
| Sputum - Xpert MTB/RIF Ultra | 998 | 95% (90-98%)<br>126/132 | 100% (96-100%)<br>90/90 | 85% (70-93%)<br>33/39 | 97% (96-98%)<br>842/866 |
| Sputum swab - MiniDock MTB | 996 | 86% (79-91%)<br>114/132 | 96% (89-98%)<br>86/90 | 67% (51-79%)<br>26/39 | 98% (96-99%)<br>844/864 |
| Tongue swab - MiniDock MTB | 997 | 76% (68-82%)<br>100/132 | 90% (82-95%)<br>81/90 | 44% (29-59%)<br>17/39 | 99% (98-99%)<br>856/865 |

\*3 with missing smear results; \*\*2 invalid sputum swab results, 1 invalid tongue swab result

As compared to sputum on Ultra, positive percent agreement of sputum swabs on the MiniDock MTB was 86% (95% CI 78-91%) at health facilities and 58% (95% CI, 45-70%) in community ACF, with poorer agreement in participants with sputum Ultra results of very low and trace (Table 3). Positive percent agreement of tongue swabs was 75% (95% CI, 66-83%) at health facilities and 42% (95% CI, 30-55%) in the community.

**Table 3.** Diagnostic agreement of tongue swabs and sputum swabs on MiniDock MTB with sputum on Xpert MTB/RIF Ultra, by result grade and setting.

|  | Positive percent agreement |  |  |  | Negative percent agreement |  |
| --- | --- | --- | --- | --- | --- | --- |
|  | Health facilities |  | Active case finding |  |  |  |
|  | % (95% CI) | n/N | % (95% CI) | n/N | % (95% CI) | n/N |
| **Sputum swabs (n= 1,093)** |  |  |  |  |  |  |
| Overall | 86% (78-91%) | 91/106 | 58% (45-70%) | 33/57 | 98.3% (97-99%) | 914/930 |
| High | 93% (83-97%) | 51/55 | 100% (61-100%) | 6/6 |  |  |
| Medium | 100% (82-100%) | 17/17 | 100% (61-100%) | 6/6 |  |  |
| Low | 94% (72-99%) | 15/16 | 83% (61-94%) | 15/18 |  |  |
| Very low | 75% (30-95%) | 3/4 | 27% (11-52%) | 4/15 |  |  |
| Trace | 36% (16-61%) | 5/14 | 17% (3-49%) | 2/12 |  |  |
| Total, minus Trace | 94% (87-97%) | 86/92 | 69% (54-80%) | 31/45 |  |  |
| **Tongue swabs (n=1,094)** |  |  |  |  |  |  |
| Overall | 75% (66-83%) | 80/106 | 42% (30-55%) | 24/57 | 99.5% (99-100%) | 926/931 |
| High | 95% (85-98%) | 52/55 | 83% (44-97%) | 5/6 |  |  |
| Medium | 76% (53-90%) | 13/17 | 100% (61-100%) | 6/6 |  |  |
| Low | 75% (51-90%) | 12/16 | 67% (44-84%) | 12/18 |  |  |
| Very low | 50% (15-85%) | 2/4 | 7% (1-30%) | 1/15 |  |  |
| Trace | 7% (1-31%) | 1/14 | 0% (0-25%) | 0/12 |  |  |

### MTB Ultima diagnostic accuracy

Among 766 MTB Ultima results on sputum swabs, overall sensitivity was 84% (95% CI, 76-89%) and sensitivity among smear-negative specimens was 58% (95% CI, 64-91%), as compared to 95% (95% CI, 89-98%) overall and 81% (95% CI, 64-91%) for smear-negative specimens on sputum Ultra (Table 4).

**Table 4.**
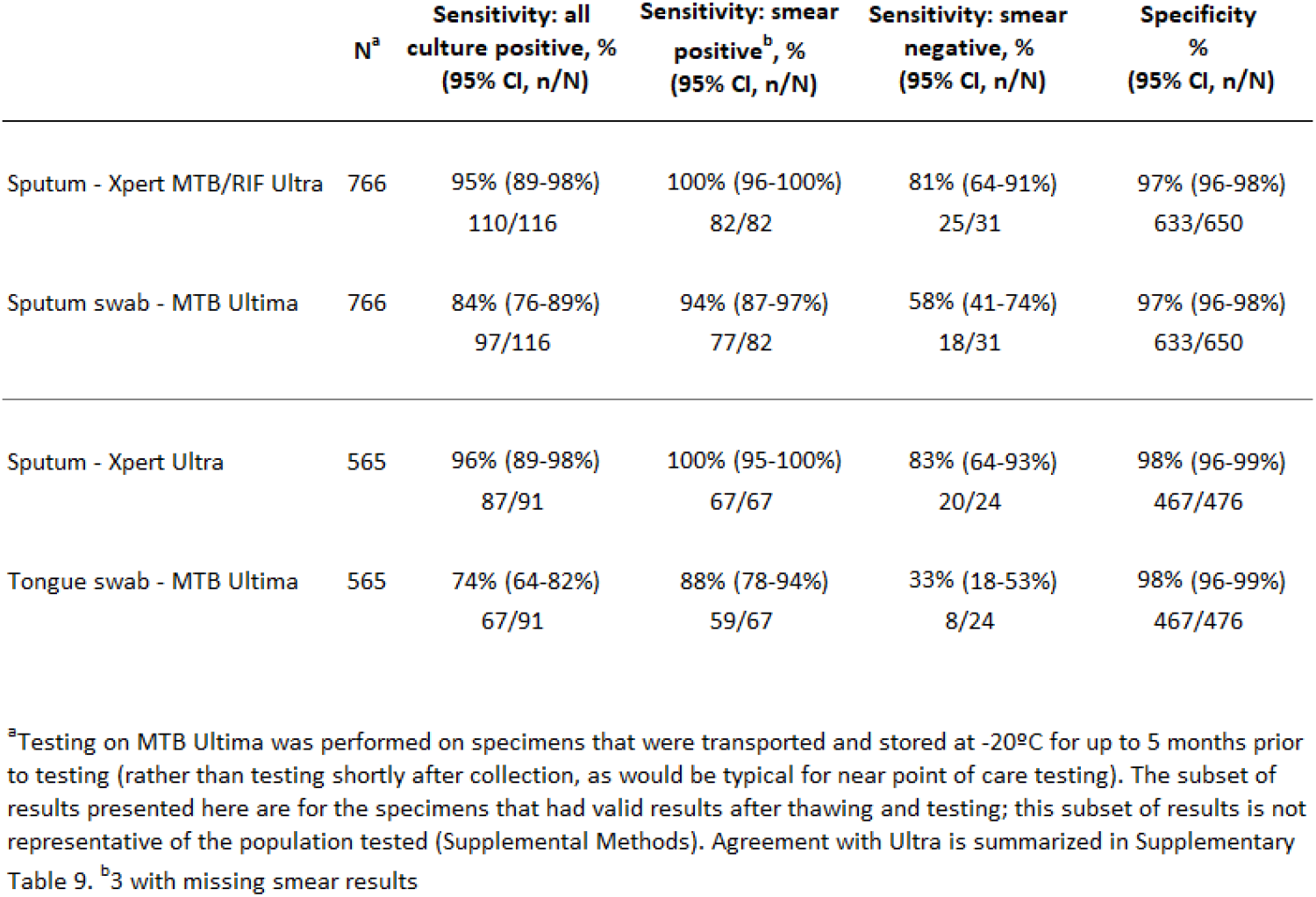
Diagnostic accuracy of sputum swabs and tongue swabs on MTB Ultima and sputum on Xpert MTB/RIF Ultra for detection of TB, as compared with the reference standard of TB culture.

|  | N<sup>a</sup> | Sensitivity: all culture positive, %<br>(95% CI, n/N) | Sensitivity: smear positive<sup>b</sup>, %<br>(95% CI, n/N) | Sensitivity: smear negative, %<br>(95% CI, n/N) | Specificity %<br>(95% CI, n/N) |
| --- | --- | --- | --- | --- | --- |
| Sputum - Xpert MTB/RIF Ultra | 766 | 95% (89-98%)<br>110/116 | 100% (96-100%)<br>82/82 | 81% (64-91%)<br>25/31 | 97% (96-98%)<br>633/650 |
| Sputum swab - MTB Ultima | 766 | 84% (76-89%)<br>97/116 | 94% (87-97%)<br>77/82 | 58% (41-74%)<br>18/31 | 97% (96-98%)<br>633/650 |
| Sputum - Xpert Ultra | 565 | 96% (89-98%)<br>87/91 | 100% (95-100%)<br>67/67 | 83% (64-93%)<br>20/24 | 98% (96-99%)<br>467/476 |
| Tongue swab - MTB Ultima | 565 | 74% (64-82%)<br>67/91 | 88% (78-94%)<br>59/67 | 33% (18-53%)<br>8/24 | 98% (96-99%)<br>467/476 |

<sup>a</sup>Testing on MTB Ultima was performed on specimens that were transported and stored at -20°C for up to 5 months prior to testing (rather than testing shortly after collection, as would be typical for near point of care testing). The subset of results presented here are for the specimens that had valid results after thawing and testing; this subset of results is not representative of the population tested (Supplemental Methods). Agreement with Ultra is summarized in Supplementary Table 9. <sup>b</sup>3 with missing smear results

Among 565 MTB Ultima results on tongue swabs, overall sensitivity was 74% (95% CI, 64-82%) and sensitivity among smear-negative specimens was 33% (95% CI, 18-53%), as compared to 96% (95% CI, 89-98%) overall and 83% (95% CI, 64-93%) for smear-negative specimens on sputum Ultra.

### Tongue swab collection self vs. healthcare worker

On tongue swabs tested on MiniDock TB, sensitivity of TB detection among people who elected supervised self-collection was 75% (84/112, 95% CI, 66-82%), similar to the sensitivity of 80% (16/20, 95% CI, 58-92%) as among those with healthcare worker collected tongue swabs (Supplemental Table 6). For tongue swabs tested on the Ultima, sensitivity for self-collection was 73% (60/82, 95% CI 63-82%), similar to 78% (7/9, 95% CI, 45-94%) for healthcare worker collected (Supplemental Table 8).

### Test success rate, time to result and operating temperature

On the MiniDock MTB, the initial test invalid rate for sputum swabs was 1.0% (11/1,096 tests) and for tongue swabs was 0.9% (10/1,096 tests); on repeat testing 2/11 and 1/10, respectively, were invalid (Supplemental Table S2).

On the MiniDock MTB, the instrument test time to generate negative results is 25 minutes; for positive results the average time to result was 13.4 minutes (SD= 4.0 minutes) for sputum swabs and 14.3 minutes (SD=4.3 minutes) for tongue swabs (Supplemental Table S3).

At ACF events, the MiniDock MTB test was routinely performed at temperatures of >35ºC (Supplemental Table S4).

## Discussion

We report a diagnostic evaluation of MTB Ultima and MiniDock MTB, including individuals recruited in community settings as well as at health facilities. Our results demonstrated performance of both assays to be robust, with swabbed sputum similar to Ultra among individuals with higher semiquantitative results on Ultra, but with poorer performance among individuals with lower semiquantitative results. In this population, MiniDock MTB met the WHO TPP thresholds for both sputum and non-sputum based near point of care tests, while MTB Ultima was within a percentage point of the targets[24]. Our results are similar to the only other early study in health facilities for both the diagnostic performance of both the assays and the low rates of test failure for MiniDock MTB[19].

The performance of both assays was less sensitive on tongue swabs in general and among individuals with lower semiquantitative results on Ultra. The differences across semiquantitative grades on Ultra may be important based on the population being tested. Although a small sample, our results suggest that people with TB identified in community settings may have lower levels of TB bacilli and a different semiquantitative results profile than those identified in health facilities[27]. Nevertheless, since most TB programs in high burden countries still test large numbers of people with microscopy, an improvement in performance compared to microscopy coupled with increased coverage at point of care could greatly enhance coverage.

Overall, staff running the tests reported both platforms were easy to use with minimal training, and workflows were straightforward enough to be done at a health facility with basic staffing and electricity. The Pluslife platform was also used directly in the field at ambient temperatures between 35-40° centigrade (Supplemental Table 4), with a solar power supply for the Thermomixer. In its current version, the MiniDock indicates results as positive or negative without a display on the instrument (with full results available if connected to an ancillary computer); however, future versions may facilitate automated and verifiable results and reporting through a smartphone (communication with manufacturer).

The Pluslife MiniDock TB assay has the advantage of running on an extremely low cost instrument (less than $200 each for the MiniDock MTB and Thermomixer), with tests that are already much lower cost than currently available NAATs (MiniDock TB <$4 vs TB LAMP $6 when optimally batched, Truenat $8, Ultra $8[28][19]) and may be even less expensive when supplied at higher volumes. The platform is >20 times less expensive than either GeneXpert and Truelab instruments and >8 times less expensive than the TB LAMP instrument. However, a major disadvantage of MiniDock MTB platform as compared to the existing automated NAAT platforms is that no rifampin resistance testing is possible. This disadvantage is shared by the TB LAMP platform and has contributed to slow uptake of this technology, due in part to the need to provide subsequent rifampin-resistance testing on a separate platform[5].

The MTB Ultima assay is performed with the new Truelyse instrument, for rapid lysis of swab specimens (90 seconds), followed by TB detection on the Truelab instrument. The Truelab instrument is already part of the platform (together with the Trueprep instrument for DNA extraction) used for testing the Truenat MTB Plus assay, which has been recommended by the WHO since 2020, with the Truenat MTB-RIF assay for detection of rifampin resistance. The Truelab is a real-time PCR instrument available for $10,000 and above[28]. The platform is portable and battery-powered, with demonstrated good performance for TB and rifampin resistance detection in peripheral and community settings.[9] While the new MTB Ultima assay has the advantage that follow-up rifampin resistance testing can be performed with the same Truelab instrument, the high cost of this instrument (at least 5 times the WHO TPP capital cost target of $2,000)[24] may represent a significant barrier for scale-up and decentralization of this new assay.

The ability of both platforms to test oral swabs presents both opportunities and implementation questions. Diagnostic performance of tongue swabs collected by participants under supervision and by healthcare workers was similar in this study (Supplementary Tables 6 and 8). Previous studies have documented large losses in the diagnostic cascade related to sputum collection and testing that an easy to use tongue swab test could potentially address[29–31]. Measuring both accuracy as well as yield with non-sputum tests is highlighted in recent publications and forms part of the updated WHO diagnostic TPP[17,32]. Another early study evaluating the same swab-based testing platforms found similar diagnostic yield between tongue swab and swabbed sputum across four countries[33], although only 85% of participants overall provided sputum. In our study, 99% of individuals provided sputum, including in community settings, aided by video coaching and/or use of Lung Flute ECO; similar high sputum collection rates have been documented in other work[34,35]. Performance of tongue swabs was lower than swabbed sputum, as expected[36,37], and during programmatic implementation it will be important to highlight the importance of obtaining sputum when possible to improve detection. The impact of decentralizing molecular testing capacity on TB diagnosis, molecular testing coverage and time to treatment are important questions to address in larger future studies.

Another approach to facilitate greater access to rapid diagnostic testing is with pooled testing. Several studies of pooled testing on Ultra have shown per person testing costs reduced by 30-60% with minimal loss in performance in diverse populations[38–40]. More evidence is needed to guide implementers about when to use higher sensitivity tests (e.g. pooled or individual Ultra) versus easier to implement tests (e.g. MiniDock MTB) to improve both access to testing and detection of TB in different populations, including cost considerations.

Our study has several limitations. We performed culture on one sputum per participant rather than on two or more sputum specimens as preferred for better detection of TB. In addition, culture was conducted on different a specimen than index testing[25]. MTB Ultima was not tested on fresh samples, but instead after sample transport and storage; this likely contributed to the high rate of invalid results on this assay; as the test is designed for use at or near point of care, this is not expected to be a concern in program use. While we included participants recruited in the community and people living with HIV, future work should include larger sample sizes and other populations like children.

## Conclusion

MTB Ultima and MiniDock MTB swab testing platforms bring the potential for TB diagnosis nearer to point-of-care. The platforms are robust, have easy to use workflows, and have clear performance improvements over smear microscopy. In certain populations, these tests have similar performance to existing low complexity NAAT platforms, and the MiniDock MTB has a much lower test and equipment cost. Future studies are needed to investigate performance in different populations, especially at lower levels of care, as well as how best to integrate swab-testing in laboratory networks, how swab testing is used and received by both people with presumptive TB and health care providers, and cost effectiveness.

## Data Availability

All data produced in the present work are contained in the manuscript

## Notes

## Acknowledgments

We thank all participants for their contribution to this work. We gratefully acknowledge all the contributions of the members of the RAPID TB Consortium: Nina Lubeka (Bonaberi Baptist Hospital, Bonaberi, Cameroon); Armand Koudjou (Bafoussam Baptist Hospital, Bafoussam, Cameroon); Guy Zero Molesa (Mboppi Baptist Hospital, Mboppi Cameroon); Joel Wepngong Tabah, Ngowo Eyambe Lydia (Mutengene Baptist Hospital, Mutengene, Cameroon); Christabelle Ewane (Bamenda Regional Hospital): Nguifu Kelly Ngwafung, Angela Neh, Ambo Noeline Niba, Kolla Magang Nelly Celestine, Bechesi Carine, Boum Delphine Agnes Gaetan, Ngu Amanda Mambo, Yinkfu Marcel Ndamnsah, Chifu Nadege Bonkar, Asonganyi Etiendem, Munsi Valerie Nformi, Hella Adzoyo Reine, Elimbi Neline Yameni, Ahmadou Djenabou, Maiyanpa Moksala Josephine, Diana Gladys Kolieghu, Nchu-Nfor Evangeline, Iya A Nyam Aicha, Laban Rhoda Bongshe, Ngapgue Sobdong Dominique Josiane, Tsimene Tamessuing Sandrine, Gingir Beatrice Ndemnwi, Amaya Alhadji Wakar, Souk-ino Wappou Ferdinand, Guidzavai Koliye Paul, Naindouba Beninga Herve Steve, Eban Odi Luisa Nang, Tchindo Dargeo, Edzengte Pascal Steve Landry, Elisée Avaikdepainani, Njikam Daouda Momgbet, Fonyuy Gilford, Mbah Romarick, Baiguerel Erika Myriam (Center for Health Promotion and Research)

## Data sharing statement

Individual participant data available in supplementary material.

