## Supplementary Materials for "Diagnostic performance of the Pluslife MiniDock MTB and Molbio MTB Ultima assays to detect tuberculosis from tongue and sputum swabs among outpatients and in active case finding in Cameroon"

### 1. Supplementary methods

#### a. Randomization of tongue swab testing order

Sequences were assigned in randomly permuted blocks of 50, stratified by site, by a person not involved in the interventions, using a random sequence generator software and sealed in sequentially numbered opaque envelopes. At the testing lab, the technician opened the envelope with the appropriate participant number and placed labels on the storage tubes containing each tongue swab with the name of the index test to be performed based on tongue swab collection order (e.g. specimen # 1-PlusLife/ specimen #2-Ultima or specimen # 1-Ultima/ specimen #2-PlusLife).

#### b. MTB Ultima testing on stored specimens

Although all sputum swabs and tongue swabs collected for testing on Ultima were tested, there were many invalid results. The high rate of invalids may be attributable to the amount of time that the specimens were stored at ambient temperatures after collection and/or during subsequent storage at -20ºC for up to five months before testing, followed by thawing. The internal control of the assay may degrade under these storage conditions. Invalid results were more common in specimens collected in the community; these specimens were collected at higher temperatures (see Supplementary Table 4) and were transported to the reference lab prior to storage, potentially contributing to the higher invalid rate.

#### c. TB culture testing

For the microbiological reference standard (MRS), individuals were considered positive for TB if any culture result was positive. At the reference laboratory, sputum specimens of ≥2 mL were initially homogenized by vortexing the specimens with 3mm sterile glass beads for 20 min. Then, the homogenized sputum was aliquoted for culture processing (2 mL) and any excess was stored in 0.5mL aliquots for additional testing as needed. Sputum was processed using the N-acetyl-L-cysteine–NaOH (NALC-NaOH) method for TB culture.[21]. After centrifugation, the pellet was resuspended in the same volume as the starting sputum (2 mL) of phosphate-buffered saline (PBS) buffer; then, 0.5 mL was inoculated on mycobacterial growth indicator tubes (MGIT; BD Diagnostic Systems, Sparks, MD, USA) using the Bactec MGIT 960 system, and 0.2 mL was inoculated on Lowenstein-Jensen media. Cultures positive for acid- fast bacilli were tested for *M. tuberculosis* complex by MPT64 antigen detection (Standard Diagnostics, Republic of Korea). Isolates that were AFB positive and negative by MPT64 antigen testing were tested for mycobacterial identification using the GenoType Mycobacterium CM and AS line probe assays (Hain Lifescience, Germany) according to the manufacturer's instructions.

### 2. Supplemental Tables

#### Table S1. Testing locations for index, comparator, and reference tests, by enrollment site

| Enrollment site | Affiliated lab(s) for index and comparator testing | MiniDock MTB test location | MTB Ultima test location[2] | Smear microscopy[3] test location | Ultra test location | TB culture test location[4] |
|---|---|---|---|---|---|---|
| **Health facility 1** | Reference Lab 1 | Ref Lab 1 | Ref Lab 1 | Ref Lab 1 | Ref Lab 1 | Ref Lab 1 |
| **Health facility 2** | Xpert Lab 1 | XP Lab 1 | Ref Lab 1 | XP Lab 1 | XP Lab 1 | Ref Lab 1 |
| **Health facility 3** | Xpert Lab 1 | XP Lab 1 | Ref Lab 1 | XP Lab 1 | XP Lab 1 | Ref Lab 1 |
| **Health facility 4** | Microscopy Lab 1, Ref Lab 1 | Ref Lab 1 | Ref Lab 1 | Mic Lab 1 | Ref Lab 1 | Ref Lab 1 |
| **Health facility 5** | Ref Lab 1 | Ref Lab 1 | Ref Lab 1 | Ref Lab 1 | Ref Lab 1 | Ref Lab 1 |
| **Community events** | Xpert Lab 2 or 3 | Community event[1] | Ref Lab 1 | Ref Lab 1 | XP Lab 2 or 3 | Ref Lab 1 |

[1] MiniDock MTB testing was performed onsite at the community event

[2] For all MTB Ultima testing, tongue and sputum samples were collected at the various enrollment sites, then transported and stored at the reference lab prior to testing when reagents were available

[3] Ziehl-Neelsen microscopy was performed at Microscopy Lab 1 and Xpert Lab 1; auramine fluorescence microscopy was performed at Ref Lab 1

[4] TB culture was performed at the Tuberculosis Reference Laboratory Bamenda, which is accredited in accordance with the recognized International Standard ISO 15189:2022 (SANAS Accredited Medical Laboratory, no. M0593).

**Table S2. Test success rate, MiniDock MTB, by swab type**

| | Overall n/N (%) |
|---|---|
| **MiniDock MTB sputum swab** | |
| Tongue swab initial invalid result | 11/1093 (1.0%) |
| Repeat sputum swab result as invalid | 2/11 (18.2%) |
| **MiniDock MTB tongue swab** | |
| Tongue swab initial invalid result | 10/1096 (0.9%) |
| Repeat sputum swab result as invalid | 1/10 (10.0%) |

**Table S3. Time to result of MTB positive sputum and tongue swabs on the MiniDock MTB instrument, overall and by Ultra sputum semi-quantitative grade**

| | Sputum swab | | | Tongue swab | | |
|---|---|---|---|---|---|---|
| | Number | Average time to results, minutes | SD, minutes | Number | Average time to results, minutes | SD, minutes |
| **Overall*** | 129 | 14.6 | 5.1 | 99 | 15.1 | 4.7 |
| Ultra MTB detected, grade | | | | | | |
| High | 55 | 11.7 | 0.9 | 53 | 14.0 | 3.7 |
| Medium | 19 | 11.9 | 0.6 | 17 | 14.1 | 3.1 |
| Low | 28 | 14.2 | 4.1 | 22 | 16.6 | 5.4 |
| Very Low | 5 | 22.9 | 4.7 | 2 | 21.3 | 5.3 |
| Trace | 6 | 21.4 | 5.7 | 0 | | |
| Ultra MTB Not Detected | 16 | 23.4 | 3.6 | 5 | 22.3 | 6.0 |

*11 positive sputum swab results with no time recorded; 10 positive tongue swab results with no time recorded

**All swabs with MiniDock MTB negative results had a test time of 25 minutes (not included in this table)

**Table S4. Ambient temperature measured at the time of swab testing with the MiniDock MTB device, by setting and test result**

| | Total | | | Active Case Finding | | | Health Facilities | | |
|---|---|---|---|---|---|---|---|---|---|
| | N | Average (◦C) | Range (◦C) | N | Average (◦C) | Range (◦C) | N | Average (◦C) | Range (◦C) |
| **Sputum swab** | | | | | | | | | |
| **Overall** | **982** | **29.0** | **[13 - 48.4]** | **377** | **35.4** | **[23 - 48.4]** | **605** | **24.6** | **[13 - 35]** |
| By result | | | | | | | | | |
| Negative | 878 | 29.1 | [13 - 48.4] | 347 | 35.4 | [23 - 48.4] | 531 | 24.6 | [13 - 35] |
| Positive | 99 | 27.9 | [17 - 43.7] | 27 | 34.8 | [23 - 43.7] | 72 | 24.9 | [17 - 32] |
| Invalid | 5 | 33.3 | [26.2 - 40.4] | 3 | 37.9 | [36.3 - 40.4] | 2 | 26.5 | [26.2 - 26.8] |
| **Tongue swab** | | | | | | | | | |
| **Overall** | **992** | **29.5** | **[17 - 44.4]** | **377** | **36.5** | **[21.7 - 44.4]** | **615** | **24.7** | **[17 - 35]** |
| By result | | | | | | | | | |
| Negative | 860 | 29.6 | [17 - 44.4] | 336 | 36.7 | [21.7 - 44.4] | 524 | 24.7 | [17 - 35] |
| Positive | 129 | 28.4 | [17 - 44.4] | 41 | 35.3 | [24.7 - 44.4] | 88 | 24.6 | [17 - 30] |
| Invalid | 3 | 23.3 | [17 - 27] | 0 | | | 3 | 23.3 | [17 - 27] |

**Table S5. Diagnostic accuracy of sputum swabs on the MiniDock MTB assay for detection of TB, as compared with the reference standard of TB culture, by sub-group**

| | **Sensitivity % (95% CI, n/N)** | **Specificity % (95% CI, n/N)** |
|---|---|---|
| Overall* | 86% (79-91%) 114/132 | 97% (96-98%) 842/866 |
| People living with HIV | 88% (64-97%) 14/16 | 97% (93-99%) 167/172 |
| People without HIV** | 86% (79-91%) 94/109 | 98% (96-99%) 597/612 |
| People enrolled at health faciltiies | 87% (79-92%) 87/100 | 98% (97-99%) 561/571 |
| People enrolled at ACF events | 84% (68-93%) 27/32 | 97% (94-98%) 283/293 |

* 2 with invalid MiniDock MTB sputum swab results

** not including 101 people with HIV status unknown

**Table S6. Diagnostic accuracy of tongue swabs on the MiniDock MTB assay for detection of TB, as compared with the reference standard of TB culture, by sub-group**

| | Sensitivity % (95% CI, n/N) | Specificity % (95% CI, n/N) |
|---|---|---|
| Overall* | 76% (68-82%) 100/132 | 99% (98-99%) 856/865 |
| People living with HIV** | 56% (33-77%) 9/16 | 100% (98-100%) 173/173 |
| People without HIV** | 80% (71-86%) 87/109 | 99% (98-99%) 606/613 |
| People enrolled at health faciltiies | 80% (71-87%) 80/100 | 100% (99-100%) 572/573 |
| People enrolled at ACF events | 63% (45-77%) 20/32 | 97% (95-99%) 284/292 |
| People with supervised self-collected swab | 75% (66-82%) 84/112 | 99% (98-99%) 714/722 |
| People with healthcare worker collected swab | 80% (58-92%) 16/20 | 99% (96-100%) 142/143 |

* 1 with invalid MiniDock MTB tongue swab result

** not including 101 people with HIV status unknown

**Table S7. Diagnostic accuracy of sputum swabs on the MTB Ultima assay for detection of TB, as compared with the reference standard of TB culture, by sub-group**

| | **Sensitivity % (95% CI, n/N)** | **Specificity % (95% CI, n/N)** |
|---|---|---|
| Overall | 84% (76-89%) 97/116 | 97% (96-98%) 633/650 |
| People living with HIV | 79% (52-92%) 11/14 | 99% (96-100%) 157/159 |
| People without HIV** | 85% (76-90%) 83/98 | 97% (95-98%) 449/462 |

** not including people with HIV status unknown

**Table S8. Diagnostic accuracy of tongue swabs on the MTB Ultima assay for detection of TB, as compared with the reference standard of TB culture, by sub-group**

| | Sensitivity % (95% CI, n/N) | Specificity % (95% CI, n/N) |
|---|---|---|
| Overall | 74% (64-82%)<br>67/91 | 98% (96-98%)<br>467/476 |
| People living with HIV** | 55% (28-79%)<br>6/11 | 98% (98-100%)<br>129/131 |
| People without HIV** | 76% (65-84%)<br>60/79 | 98% (98-99%)<br>326/333 |
| People with supervised self-collected swab | 73% (63-82%)<br>60/82 | 98% (98-99%)<br>399/407 |
| People with healthcare worker collected swab | 78% (45-94%)<br>7/9 | 99% (96-100%)<br>68/69 |

** not including people with HIV status unknown

**Table S9. Diagnostic agreement of sputum and tongue swabs on the MTB Ultima assay for detection of TB, as compared with sputum on the Xpert MTB/RIF Ultra assay, by result grade**

| | **Positive percent agreement** | | **Negative percent agreement** | |
|---|---|---|---|---|
| | % (95% CI) | n/N | % (95% CI) | n/N |
| **Sputum swabs** (n=814) | | | | |
| MTB Detected | | | | |
| High | 96% (88-99%) | 55/57 | | |
| Medium | 90% (71-97%) | 19/21 | | |
| Low | 85% (68-94%) | 23/27 | | |
| Very low | 67% (35-89%) | 6/9 | | |
| Trace | 22% (9-45%) | 4/18 | | |
| MTB Not detected | | | 98.4% (97-99%) | 673/684 |
| **Tongue swabs** (n=605) | | | | |
| MTB Detected | | | | |
| High | 90% (78-96%) | 44/49 | | |
| Medium | 81% (57-93%) | 13/16 | | |
| Low | 52% (32-72%) | 11/21 | | |
| Very low | 25% (5-70%) | 1/4 | | |
| Trace | 0% (0-30%) | 0/9 | | |
| MTB Not detected | | | 98.6% (97-99%) | 499/506 |
